# Clinical Note Comparison and Data Retrieval Via Embedding Vectors: Model Selection, Metrics, and Convergence

**DOI:** 10.64898/2026.05.12.26352832

**Authors:** Alexandra Dahlberg, Olli Tapiola, Rami Luisto, Tuukka Puranen, Enni Sanmark, Ville Vartiainen

## Abstract

**Background:** Embedding models are an integral part of generative AI architectures, transforming text into *embedding vectors* that represent semantic content in numerical form. Despite their central role, their performance in clinical settings remains underexplored. We evaluate embedding models across two tasks: semantic difference detection in clinical texts, and data retrieval from patient records.

**Methods:** Eight models were applied to synthetic discharge summaries in English, Finnish, and Swedish. Semantic sensitivity was assessed by introducing controlled perturbations (deletion, modification, and paraphrasing) at three levels of severity; cosine similarity, and L^1^ and Euclidean distances were computed between the vectors of the original and perturbed texts. Partial vectors were compared to explore dimensionality reduction. Two models with the biggest contrast in semantic difference detection were evaluated on retrieval of relevant information from real Finnish vascular surgery records.

**Results:** Embedding vectors captured semantic differences in clinical text: content deletion and modification produced larger increases in vector distance than paraphrasing. On average, models detected the direction of semantic change correctly, but case-level performance varied considerably. Qwen3-Embedding-8B was the only model with zero directional errors, while multilingual-E5-large erred in 13.8% of cases. In data retrieval, Qwen3-Embedding-8B again outperformed multilingual-E5-large, though the margin was narrower: sufficiency scores were 3.25 vs. 3.17 out of 5 for the first query and 2.25 vs. 1.15 out of 5 for the second query. For some models, as few as 0.6-1.2% of dimensions sufficed to replicate full-vector accuracy; principal component analysis and coordinate-level analysis did not account for this finding.

**Conclusions:** Our results show that the choice of embedding model is important: performance differences between models can be large enough to determine whether clinically relevant information reaches the end user, and model weaknesses can be both task-specific and context-dependent.

## 1 Introduction

Generative artificial intelligence has become a rapidly developing framework for processing and generating natural language [1] whose applications in healthcare include clinical documentation, summarization, and decision support [2–4]. Central to many of its processes are *embedding model*s [5] which transform text into *embedding vectors* that can be compared and analysed mathematically [6]. These vectors are lists of numbers representing points in a high-dimensional coordinate space, where proximity between points aims to reflect semantic similarity between the corresponding expressions.

Many embedding models are developed predominantly using English-language general-domain data, raising concerns about their reliability in the clinical context and non-English languages [7]. These limitations are particularly consequential in multilingual healthcare settings.

This work addresses three intertwined questions: how well embedding models detect semantic changes in clinical texts, how model choice affects clinical data retrieval, and how much of the underlying model structure is actually utilised for these kinds of tasks.

## 2 Methods

### 2.1 Embedding Models

We selected eight embedding models, ensuring a mix of open-source and closed-source models with different structures and sizes.

1. **text-embedding-3-large** “OpenAI Large”) is one of the current flagship models of OpenAI. It is a closed-source model that produces 3072-dimensional embedding vectors. [8]
2. **text-embedding-ada-002** (“OpenAI Ada”) is an older model by OpenAI. It is a closed-source model with 1536-dimensional embedding vectors. [9]
3. **Nomic Embed Text V2** (“Nomic Embed V2”) is a model by Nomic AI which aims to improve performance speed and reduce memory. It is an open-source model with 768-dimensional embedding vectors. Quantization: F32. [10, 11]
4. **BGE-M3** is an embedding model by BAAI developed using data in over 170 languages. It is an open-source model with 1024-dimensional embedding vectors. Quantization: F16. [12]
5. **Qwen3-Embedding-8B** (“Qwen3-8B”) is the largest embedding model in the Qwen3 model series by Zhang, et al. [13]. It is an open-source model with 4096-dimensional embedding vectors. Quantization: Q6_K. [14]
6. **multilingual-E5-large** is the largest version of the multilingual E5 models by Wang, et al. [15]. It is an open-source model with 1024-dimensional embedding vectors. Quantization: F16. [16]
7. **EmbeddingGemma** is a model by Google which is optimized for devices with less computational power. It is an open-source model with 768-dimensional embedding vectors. Quantization: F32. [17]
8. **Arctic Embed 2.0** is a multilingual model by Snowflake. It is an open-source model with 1024-dimensional embedding vectors. Quantization: F16. [18]

### 2.2 Multilingual Synthetic Discharge Summaries

We drew 15 synthetic discharge summaries from MedBench [19], five per language across English, Swedish, and Finnish. These languages represent a range of morphological complexity and data availability. English is a high-resource language with low morphological complexity; Swedish is lower-resource with comparable morphological complexity to English; Finnish is similarly low-resource but has higher morphological complexity. Resource level influences model development, as it requires a lot of data, while morphological complexity affects retrieval, as words may appear in a multitude of forms.

To evaluate semantic difference detection, we introduced three types of controlled perturbations to each summary.

1. **Delete:** Removal of information. For example, removing a sentence or expression describing an ancillary diagnosis or medication.
2. **Modify**: Changes that do not preserve the meaning. For example, replacing words or short phrases with expressions describing the opposite.
3. **Paraphrase**: Changes that preserve the meaning. For example, replacing words with synonyms, and rewording expressions without removing information.

Each type was applied at three levels of severity: Level 1 (5 alterations), Level 2 (10 alterations), and Level 3 (15 alterations). The perturbations were crafted manually by a trilingual physician to ensure that they were comparable across languages.

Each original summary yielded 9 perturbed versions, giving 10 summaries per original and 150 in total. Average length was 306 words and 2070 characters for English, 269 and 1914 for Swedish, and 242 and 2101 for Finnish. The synthetic discharge summaries are available in the article repository.

### 2.3 Semantic Analysis Via Embedding Vectors

Average cosine similarity, L^1^ distance, and Euclidean distance for each model, language, and perturbation type were computed between the vectors of the original cases and their perturbed versions. Linear regression was used to model the perturbation effect on the metrics in each language and perturbation type. Additionally, we examined individual instances where semantic difference between texts increased but directional difference or distance between the corresponding vectors decreased for Delete and Modify. These changes were assessed between level 1 and level 2, and between level 2 and level 3.

### 2.4 Partial Vectors and Convergence Rate

We chose the largest (Qwen3-8B) and smallest (EmbeddingGemma) models to analyse if a smaller number of vector dimensions suffices for our results. For each summary and its perturbed versions, we generated embedding vectors and computed vector differences. For each difference vector and n = 1, 10, 25, 50, 100, we created 15 partial difference vectors consisting of the first n values, the n biggest absolute values, and the n smallest absolute values of the vector. We then repeated the analysis from the previous subsection for partial vectors and L^1^ values and considered how close we are to the full values for each n.

### 2.5 PCA and Coordinate Frequency

For Qwen3-8B and EmbeddingGemma, *principal component analysis* (PCA) was applied to analyse how much variance is concentrated to different directions among the vectors. Complementing this, a *coordinate frequency analysis* was conducted by recording the vector dimensions where the 100 largest changes (and therefore the largest contributions to L^1^ values) happen.

### 2.6 Data Retrieval from Real Patient Data

To study how well semantic accuracy translates to accurate data retrieval, we chose the two models showing the biggest performance contrast in difference detection: Qwen3-8B and multilingual-E5-large. We examined whether this contrast persists in data retrieval using Finnish vascular surgery notes from HUS Helsinki University Hospital. We constructed a dataset from patients with at least 300 clinical notes, and 20 patients were randomly sampled for all data retrieval tests using a fixed seed.

Each patient record was split into text chunks of 300 characters with a 25-character overlap, tracking which chunk originated from which note. Embedding vectors were generated from the chunks and the retrieval question, and cosine similarities were computed between them. The three chunks with the highest similarity, along with their corresponding notes, were selected for human evaluation.

The process was applied to each record with for both models using two questions in Finnish. The questions considered:

1. antithrombotic medication (exact question: *“Mikä on potilaan antitromboottinen lääkitys (ASA, klopidogreeli, antikoagulaatio kuten DOAC/varfariini)?”*), and
2. main vascular diagnosis and prior interventions (exact question: *“Mikä on potilaan keskeinen verisuonikirurginen sairaus (diagnoosi ja sairastunut alue/puoli) ja mitä verisuonitoimenpiteitä tai hoitoja potilaalle on tehty?”*).

The text chunks and the corresponding notes were evaluated by two blinded physicians. The materials were reviewed jointly until a consensus was reached. The texts were scored in two categories:

- Does the text answer the question? (Yes = 1, No = 0)
- How well does the text answer the question? (scale of 0-5 points)

Performance was compared using non-parametric paired tests, appropriate given the ordinal outcome scale and small sample size. For the sufficiency outcome, an exact Wilcoxon signed-rank test was used, with rank-biserial correlation as the effect size measure. Effect sizes were interpreted using conventional thresholds (0.1 small, 0.3 medium, 0.5 large) [20]. For the binary outcome, an exact McNemar’s test was applied to discordant pairs. All tests were two-sided. Descriptive statistics are mean and median with interquartile range.

### 2.7 Tools

The evaluations were done using Microsoft Visual Studio Code (VS Code, version 1.112.0; Microsoft Corporation, Redmond, WA, USA) and Prompt flow extension version 1.18.0 [21] (promptflow-tools, version 1.5.0; Microsoft Corporation, Redmond, WA, USA). Open-source models were run with Ollama version 0.13.2 [22] with the specific quantizations downloaded from Hugging Face [23]. OpenAI’s models were used through OpenAI API (San Francisco, CA, USA). Python version 3.11.9 (Python Software Foundation, Wilmington, USA) [24–27]. All scripts used are available. Real records were handled in the HUS Acamedic environment [28], in accordance with privacy requirements.

## 3 Results

### 3.1 Cosine Similarity Averages

Average cosine similarities for English summaries are presented in Figure 1.

**Figure 1.**
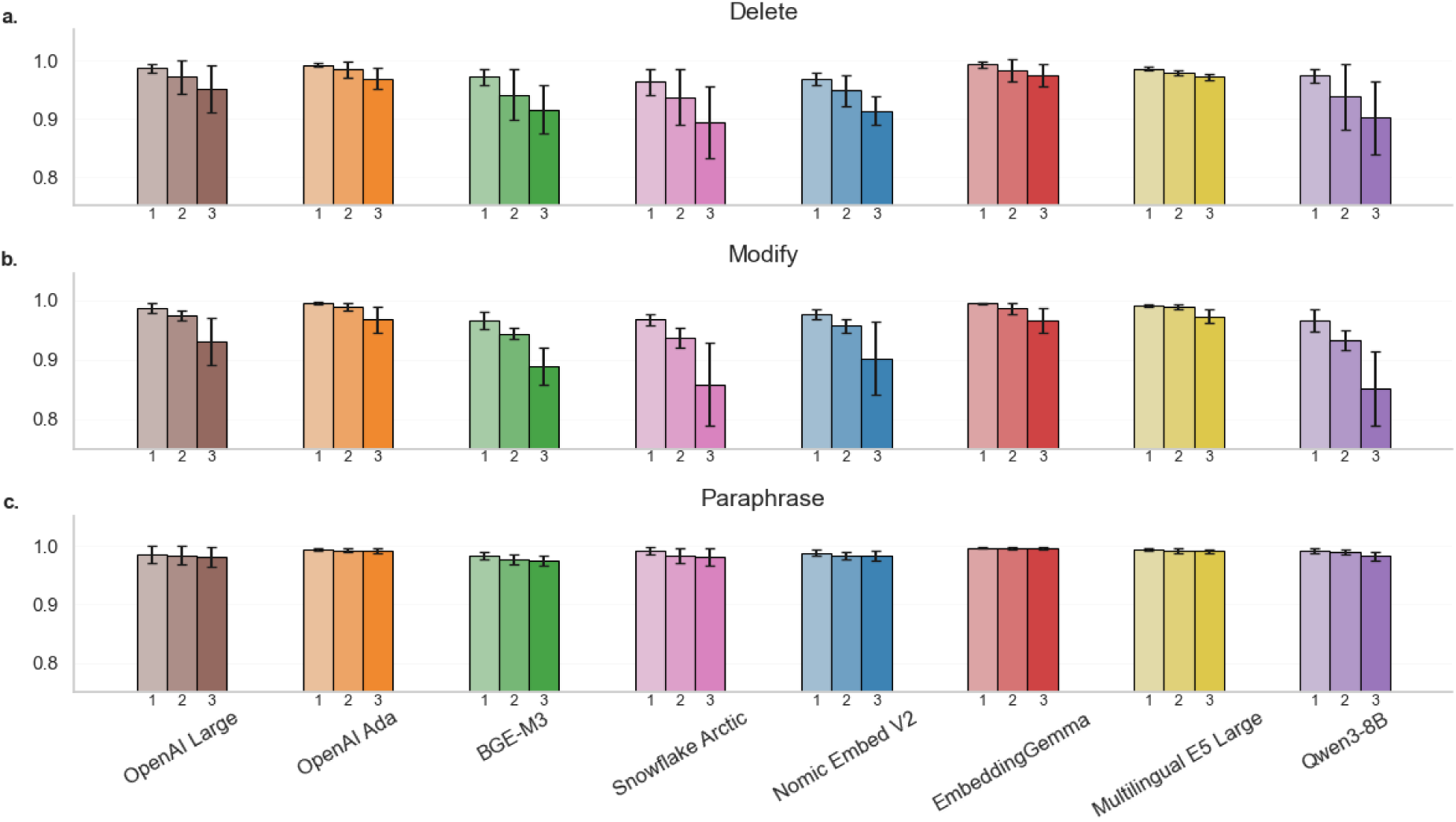
Cosine similarity between embedding vectors of English summaries. The bars show average cosine similarities over the comparisons between vectors of original English summaries and their perturbed versions of the same type (Delete, Modify, Paraphrase) and level (1 to 3). The black markers overlapping the bars represent standard deviations.

The effect of perturbation level on cosine similarity was consistent across languages, with β coefficients for Delete ranging from −0.028 to −0.032, for Modify from −0.033 to −0.038, and for Paraphrase from −0.004 to −0.006 across English, Swedish, and Finnish (Supplemental Table S1). Results for Swedish and Finnish are in Supplemental Figures S1 and S2.

### 3.2 L^1^ and Euclidean Distance Averages

BGE-M3, OpenAI Large, and Qwen3-8B showed clear differences in the cosine similarity values as the perturbation level increased. Their corresponding average L^1^ and Euclidean distance values for English summaries are presented in Figure 2.

**Figure 2.**
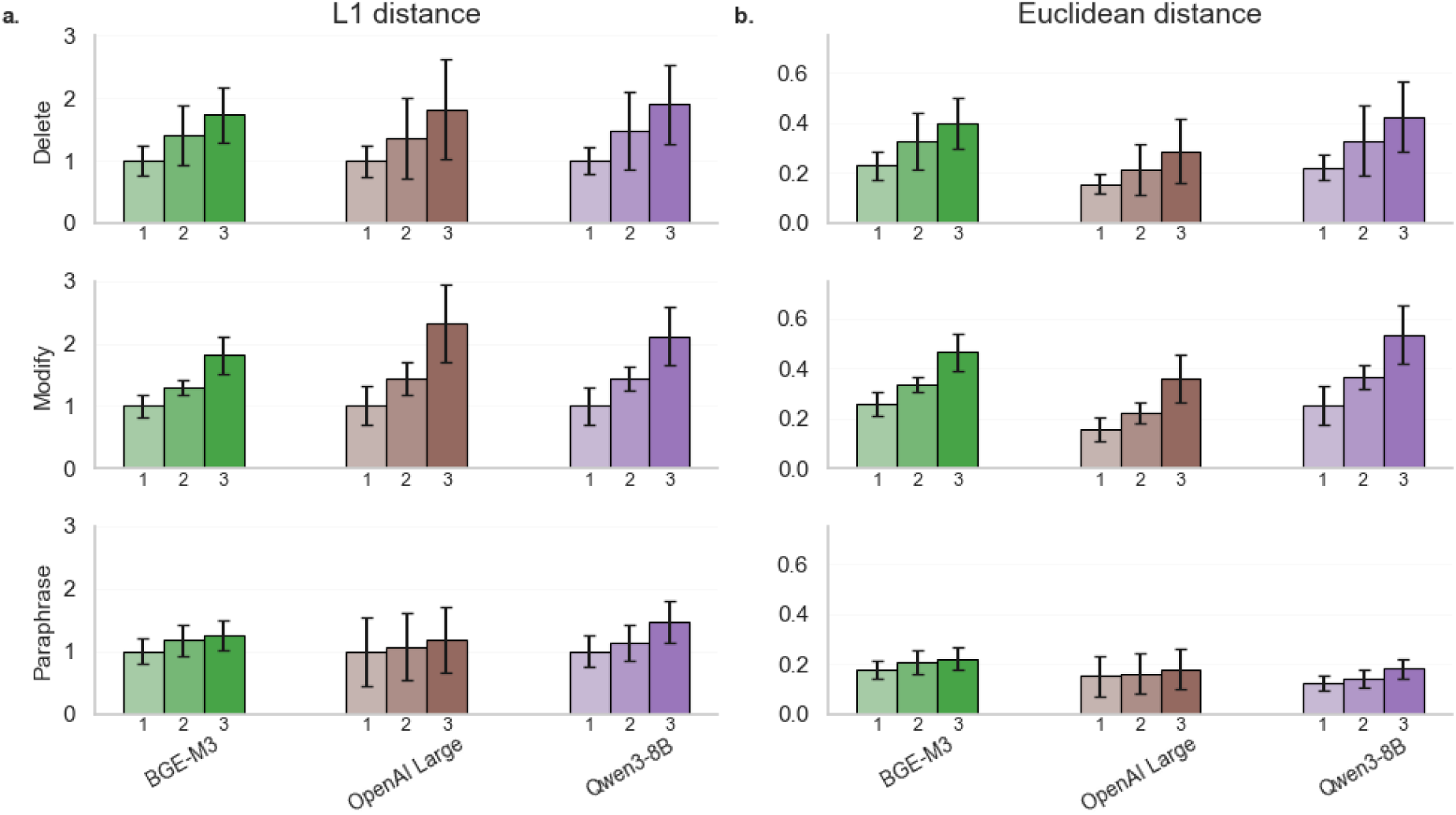
L^1^ and Euclidean distances between embedding vectors of English summaries for three models. The bars show average distances between vectors of original English summaries and their perturbed versions of the same type (Delete, Modify, Paraphrase) and level (1 to 3). The black markers overlapping the bars represent standard deviations. Since dimensionality affects L^1^ values strongly, they have been scaled so that level 1 value equals 1 to help comparability between models.

L^1^ distance β coefficients for Delete ranged from 2.141 to 2.342, for Modify from 2.308 to 2.663, and for Paraphrase from 0.532 to 0.699 across English, Swedish, and Finnish; Euclidean distance β coefficients ranged from 0.085 to 0.093 for Delete, 0.092 to 0.103 for Modify, and 0.021 to 0.028 for Paraphrase (all p ≤ 0.001; Supplemental Table S1). Results for Swedish and Finnish are in Supplemental Figures S3 and S4.

### 3.3 Cosine Similarity, L^1^, and Euclidean Distance: Individual Cases

Differences between models were more pronounced at the individual case level. For cosine similarity, Qwen3-8B was the only model with no conflicting changes, while BGE-M3 and OpenAI Large each recorded one. multilingual-E5-large showed the highest rate of conflicting changes, with 7 out of 60 changes (11.7%). When all three metrics were considered, a conflicting change in one metric was typically accompanied by a change in the other two. Total conflicting changes for each model are presented in Table 1.

**Table 1.** Number of conflicting changes per model and metric in Delete and Modify. Conflicting change was defined as a change in the wrong direction between successive perturbation levels (an increase in cosine similarity or a decrease in L¹ or Euclidean distance).

| Model | Cosine similarity | L<sup>1</sup> distance | L<sup>2</sup> distance | Total (/180) |
| --- | --- | --- | --- | --- |
| OpenAI Large | 1 | 1 | 1 | 3 |
| OpenAI ADA | 2 | 3 | 2 | 7 |
| BGE-M3 | 1 | 1 | 1 | 3 |
| Snowflake Arctic V2 | 5 | 5 | 5 | 15 |
| Nomic Embed V2 | 3 | 4 | 3 | 10 |
| EmbeddingGemma | 3 | 3 | 3 | 9 |
| multilingual-E5-large | 7 | 8 | 7 | 22 |
| Qwen3-8B | 0 | 0 | 0 | 0 |

Notably, conflicting behaviour was not distributed evenly across cases: all 9 conflicting changes for EmbeddingGemma happened in the same case for the level 2 to level 3 transition in Delete. Detailed results grouped by language and model are provided in Supplemental Tables S2, S3, and S4.

### 3.4 Partial Data and Convergence

Qwen3-8B and EmbeddingGemma were selected to explore dimensionality reduction. Scaled maximal average L^1^ values for partial difference vectors are presented in Figure 3.

**Figure 3.**
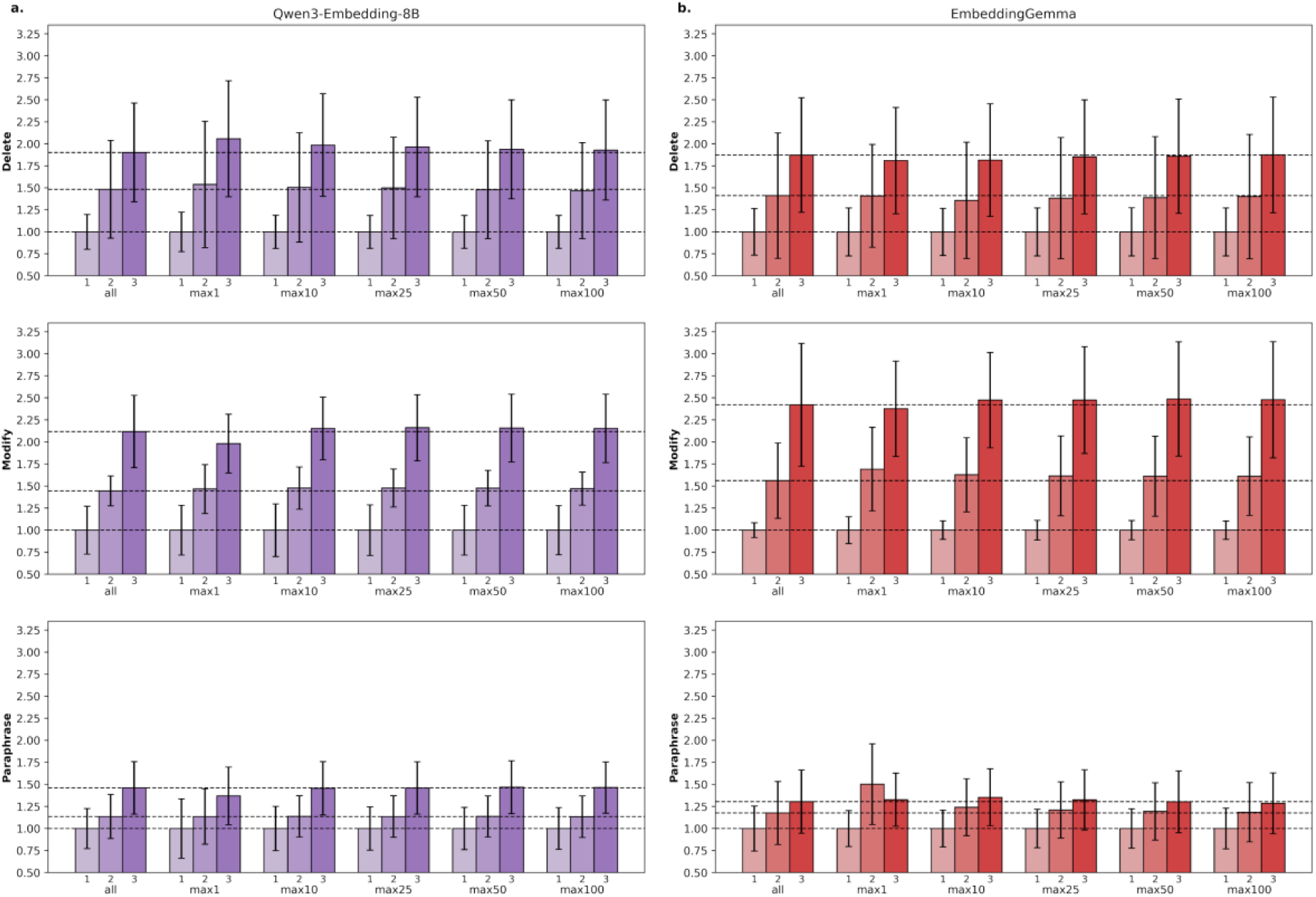
Scaled L^1^ averages for partial maximal data in English. The bars show the original average values and the corresponding values for partial difference vectors between the original and the perturbed summaries with respect to Qwen3-8B (column **a.**) and EmbeddingGemma (column **b.**) for the 1, 10, 25, 50 and 100 biggest absolute value changes. The black markers overlapping the bars represent standard deviations. In each category, the values have been divided by the corresponding level 1 value to normalize the scales between the truncation levels while preserving the relative jumps between the perturbation levels.

Scaled “partial” L^1^ averages for the biggest absolute values converged quickly towards the original scaled L^1^ values for both models. Truncation type affected convergence rates more than language. Going from the biggest first to the smallest 100 values, the differences between the scaled averages for the full and partial vectors were at most 1.7%, 5.3% and 10.8% for Qwen3-8B and 3.2%, 7.3% and 17.8% for EmbeddingGemma across all languages and perturbation types. Detailed results are provided in Supplemental Tables S5 and S6.

On the level of individual cases, Qwen3-8B required only 25 biggest values (0.6%) or 50 first values (1.2%) to reach the earlier level of zero conflicting L^1^ changes while 100 smallest values were not enough for this. For EmbeddingGemma, the earlier level was not attained in any truncation type.

### 3.5 PCA on Embedding Vectors

PCA showed a strong concentration of variance for both models. For Qwen3-8B, first two principal components accounted for 19.7% and 13.7% of the total explained variance, respectively, and it took 28 principal components to reach 95 percent of the cumulative explained variance. For EmbeddingGemma, the corresponding numbers were 21.5% and 17.0%, and 22 principal components PCA also showed a mild imbalance in variances in the original dimensions. Cumulative variances for both models are shown in Figure 4.

**Figure 4.**
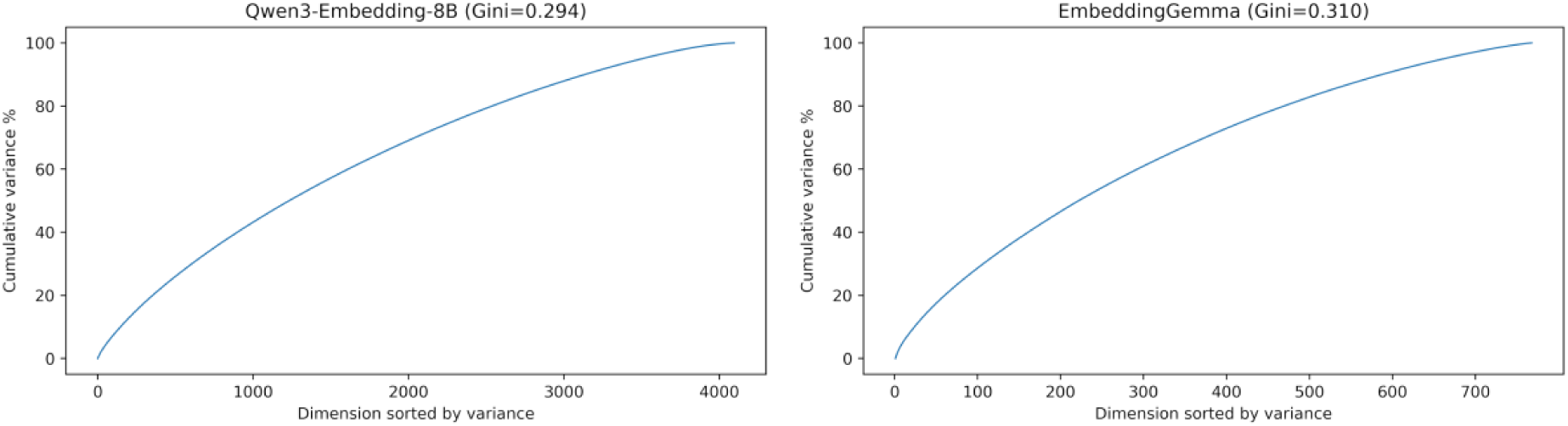
Variance accumulation for Qwen3-8B and EmbeddingGemma when dimensions are sorted by variance from biggest to smallest. The Gini coefficients for variances were 0.294 and 0.310, respectively, indicating moderate anisotropy.

For both models, variance was considerably higher than average in a handful of directions but the average change rate from biggest to smallest was quite slow.

### 3.6 Coordinate Frequencies

No coordinates appeared consistently among the 100 biggest changes for either model. Among all difference vectors for Qwen3-8B, the most frequent coordinates appeared 47 times out of a possible 135 times. For EmbeddingGemma, the number was 71. The dimensions that never appeared among the 100 biggest changes for multilingual and English texts are presented in Table 2.

**Table 2.** Dimensions that never appeared among the 100 biggest changes. We computed the difference vectors and recorded where the 100 biggest changes happened. The number of dimensions that never appeared among the 100 biggest changes increased for both models from multilingual to monolingual text collections and from all perturbation types to a single perturbation type.

| Model | Perturbation type | Language | Number of vectors | Dimensions outside top 100 (%) |
| --- | --- | --- | --- | --- |
| Qwen3-8B | All | All | 135 | 19.9 |
| Qwen3-8B | All | English | 45 | 47.7 |
| Qwen3-8B | Delete | All | 45 | 51.3 |
| Qwen3-8B | Delete | English | 15 | 76.1 |
| EmbeddingGemma | All | All | 135 | 0.4 |
| EmbeddingGemma | All | English | 45 | 4.7 |
| EmbeddingGemma | Delete | All | 45 | 4.8 |
| EmbeddingGemma | Delete | English | 15 | 26.7 |

Detailed results are provided in Supplemental Table S7.

### 3.7 Data Retrieval

Qwen3-8B and multilingual-E5-large frequently ranked different notes as the best matches: the mean number of identical retrieved notes between the models was 1.3 out of 3 for the antithrombotic therapy question, and 0.125 out of 3 for the vascular diagnosis and prior interventions question. For the vascular diagnosis question, Qwen3-8B achieved substantially higher chunk-level sufficiency scores (Wilcoxon exact test: W = 10.5, p = 0.003, rank-biserial r = 0.912), with the difference no longer significant at whole note level (p = 0.734). Human evaluation results are summarised in Figure 5.

**Figure 5.**
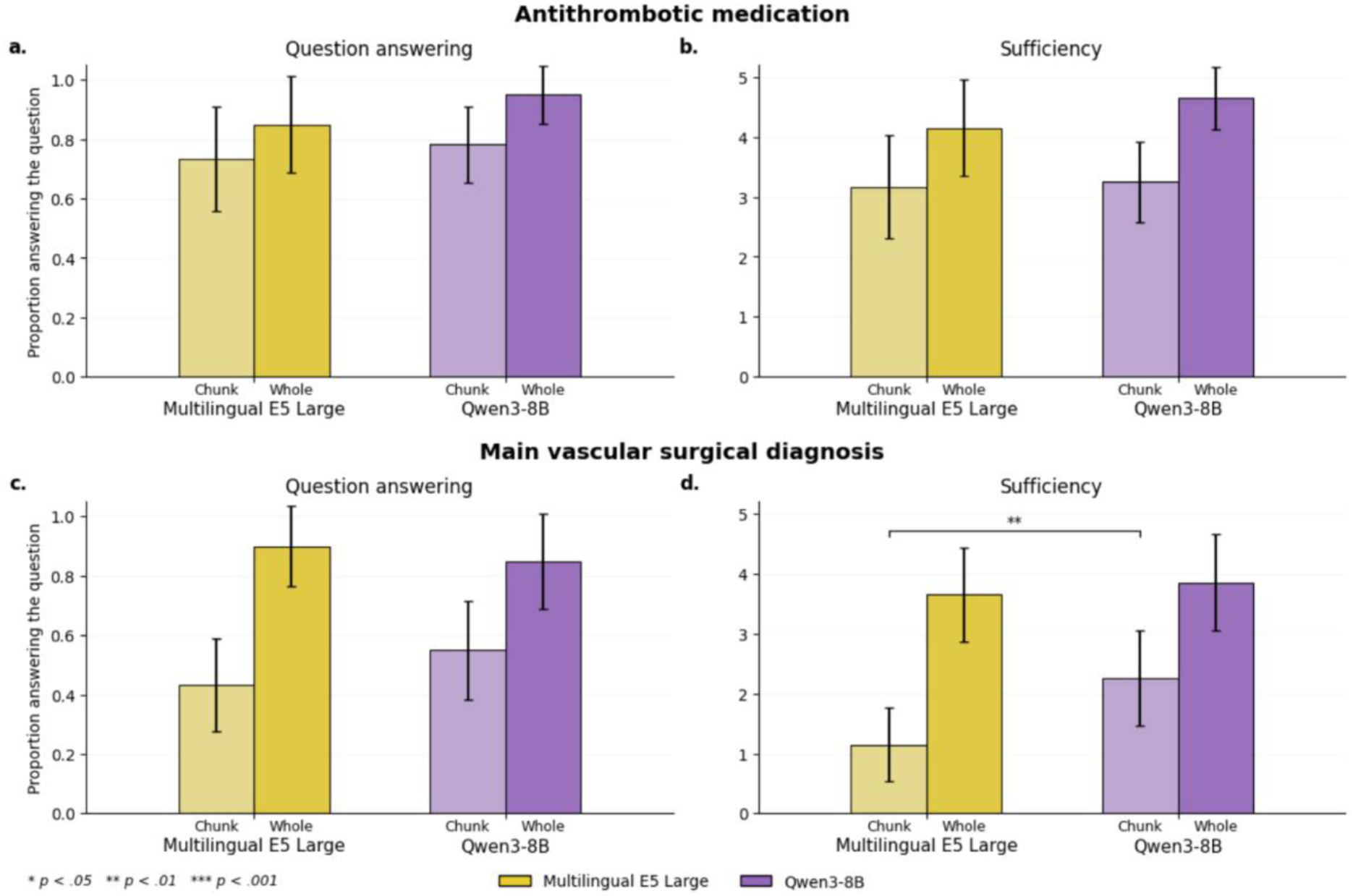
Human review results. Human evaluation results for both models across two clinical questions, with 95% confidence intervals. Panels a and b show results for the antithrombotic medication question, and panels c and d for the main vascular surgical diagnosis question. For each model, physicians rated both the highest-ranked chunk and the corresponding whole note on two criteria: whether the retrieved text answered the question (proportion, panels a and c) and the sufficiency of the information retrieved (1 to 5 scale, panels b and d).

A frequent undesired pattern was observed for Qwen3-8B: the model ranked highly short responses consisting of a single clinical term, such as “maintenance hemodialysis”. These responses did not constitute complete answers to the queries and were assigned the score 0. This pattern was observed in 6 retrieved chunks for the first question and 10 for the second.

## 4 Discussion

Average performance metrics alone were insufficient to characterise embedding model behaviour, and strong performance in one task did not guarantee equivalent performance in another. In semantic difference detection, only Qwen3-8B produced no qualitative errors across all cases, metrics, and languages, while some models that performed well on average showed high individual-level failure rates, a distinction that matters considerably in clinical settings. Language and the choice of mathematical metric had little influence on the overall pattern. For Qwen3-8B, we were able to replicate the semantic accuracy of the model with just a small fraction of the total dimensions. This is consistent with findings on the dimensionality of language models, where optimizing a small number of parameters can affect the full picture in a major way [29].

In clinical data retrieval, the performance advantage of Qwen3-8B over multilingual-E5-large declined: it was statistically significant at the chunk level for the vascular diagnosis question, diminished at the whole note level, and absent for the antithrombotic therapy question.

The near-identical regression coefficients across English, Swedish, and Finnish suggest that the models do not require language-specific adaptation, at least for the perturbation types studied here. This is broadly consistent with findings on cross-lingual transfer in multilingual embedding models, though it should be noted that evaluation standards for Scandinavian languages and Finnish remain underdeveloped [30–33].

Even though dimensionality-wise Qwen3-8B is over 5 times larger than EmbeddingGemma, Qwen3-8B reached the earlier accuracy levels with smaller number of dimensions than EmbeddingGemma in convergence analysis. This suggests that the relationship between model dimensionality and effective dimensionality is non-trivial and model-specific. PCA confirmed a concentration of variance in a small subset of directions, consistent with the well-established anisotropic nature of embedding vectors [30], which has been linked both to the concentration of semantic structures along a small number of dimensions [30] and to incorrectly detected links between unrelated texts [31]. However, the principal components did not correspond to the original coordinate directions, meaning that the convergence we observed cannot be explained by PCA alone.

Coordinate frequency analysis revealed one of the clearest language effects in the study: number of active coordinate dimensions increased considerably when the summary collection contained texts in 3 languages instead of just 1. With a single-language collection, even over 75% of dimensions remained relatively inactive. This gave us another suggestion of the context-dependent nature of embedding models, this time also from language point of view.

Despite the potential of embedding model based semantic search and retrieval-augmented generation in healthcare, such methods remain largely untapped in clinical settings [32]. Our data retrieval tests underlined one of the reasons why the use of this type of machinery requires frequent critical evaluation in context-specific tasks: Qwen3-8B often prioritized short and vascular-surgery-specific phrases above more comprehensive clinical passages. Similar phenomena related to false positives in retrieval tasks have been observed in cross-lingual settings [33]. This illustrates that embedding model behaviour may become less predictable as the context becomes specific. In addition, fixed-length chunking can split concepts into different chunks and reduce retrieval precision. Recent evaluation in clinical context has shown that adaptive chunking substantially outperforms this simpler strategy [34].

This study has limitations. Most importantly, the datasets are small: five synthetic cases as a starting point in the semantic evaluation and 20 patients with two clinical questions in the retrieval evaluation. That said, our semantic difference detection setup tested the models in three ways in three languages, and our findings were consistent across them. In addition, our retrieval setup was sufficient to reveal a context-specific weakness in Qwen3-8B. However, we compared the models only in fixed quantizations and did not study fine-tuning, which could have affected the models considerably. We also point out that the retrieval evaluation relied on consensus physician scoring without independent inter-rater reliability assessment.

## 4.1 Conclusions

Average performance metrics alone are insufficient to characterise embedding model behaviour, and strong performance in one task does not guarantee equivalent performance in another. Qwen3-8B was the only model to produce no qualitative errors across all cases in semantic difference detection, and its semantic accuracy was largely replicable with a small fraction of its total dimensions. Taken together, these findings suggest that embedding model performance is both task-specific and context-dependent, even within clinical context. These findings illustrate that embedding model evaluation must be conducted in the specific context of intended use, with attention to individual-level failures rather than aggregate metrics alone.

## 5 Declaration Statements

## 5.1 Data Availability

All data generated in this study are available within the published article or its supplementary materials. Due to data protection regulation, patient-level data cannot be shared.

## 5.2 Code Availability

The source code supporting this study is accessible at https://github.com/achdahlberg/embedding-vectors-2026.git. The tool kit is available at https://github.com/HYGenAID/Tools.git.

## 5.3 Acknowledgements

The authors gratefully acknowledge Max Gordon and the creators of the original discharge summaries included in the MedBench dataset [19]. During the preparation of this work, Claude was used (Anthropic) [35]. This tool was utilized to improve the clarity and coherence of the language, and work with some routine parts of the required programming tasks. The tool was only involved in base code editing work and was not used for any insights or exploration. After using this tool, the authors reviewed and edited the content as needed and take full responsibility for the content of the published article.

This work was supported by the Strategic Research Council at the Research Council of Finland (grant number 372591) under GAINS (“Generative AI and Digital Solutions: Enhancing Effectiveness and Productivity of Healthcare Services” and by Business Finland (1562/31/2024) through the GenAID research project. AD acknowledges Finska Läkaresällskapet, Frans Wilhelm och Waldermar von Frenckells understödsfond, and The Finnish Medical Foundation for the award of a personal grant. ES acknowledges Finska Läkaresällskapet, The Finnish Medical Foundation and, Liv och Hälsa Medical Support Association.

## 5.4 Ethics Statement

The regional healthcare authority, HUS Helsinki University Hospital approved the research permit and data use agreement (decision number HUS/355/2025). This study was based on secondary use of administrative data, in accordance with the Finnish Act on the Secondary Use of Health and Social Data (552/2019, individual patient consent or separate ethical review was not required. The study is compliant with applicable data protection regulations, including the EU General Data Protection Regulation (GDPR 2016/679).

## 5.5 Author contributions

**AD:** Conceptualisation, Data curation, Formal analysis, Investigation, Methodology, Software, Validation, Visualization, Writing - original draft. **OT:** Conceptualisation, Formal analysis, Investigation, Methodology, Software. Visualization, Writing - original draft. **RL:** Conceptualisation, Writing - review and editing. **TP:** Conceptualisation, Software, Writing - review and editing. **VV:** Funding Acquisition, Resources, Supervision, Investigation, Validation, Writing - review and editing. **ES:** Funding Acquisition, Resources, Supervision, Writing - review and editing.

## Supplemental information

**Supplemental Figure S1.**
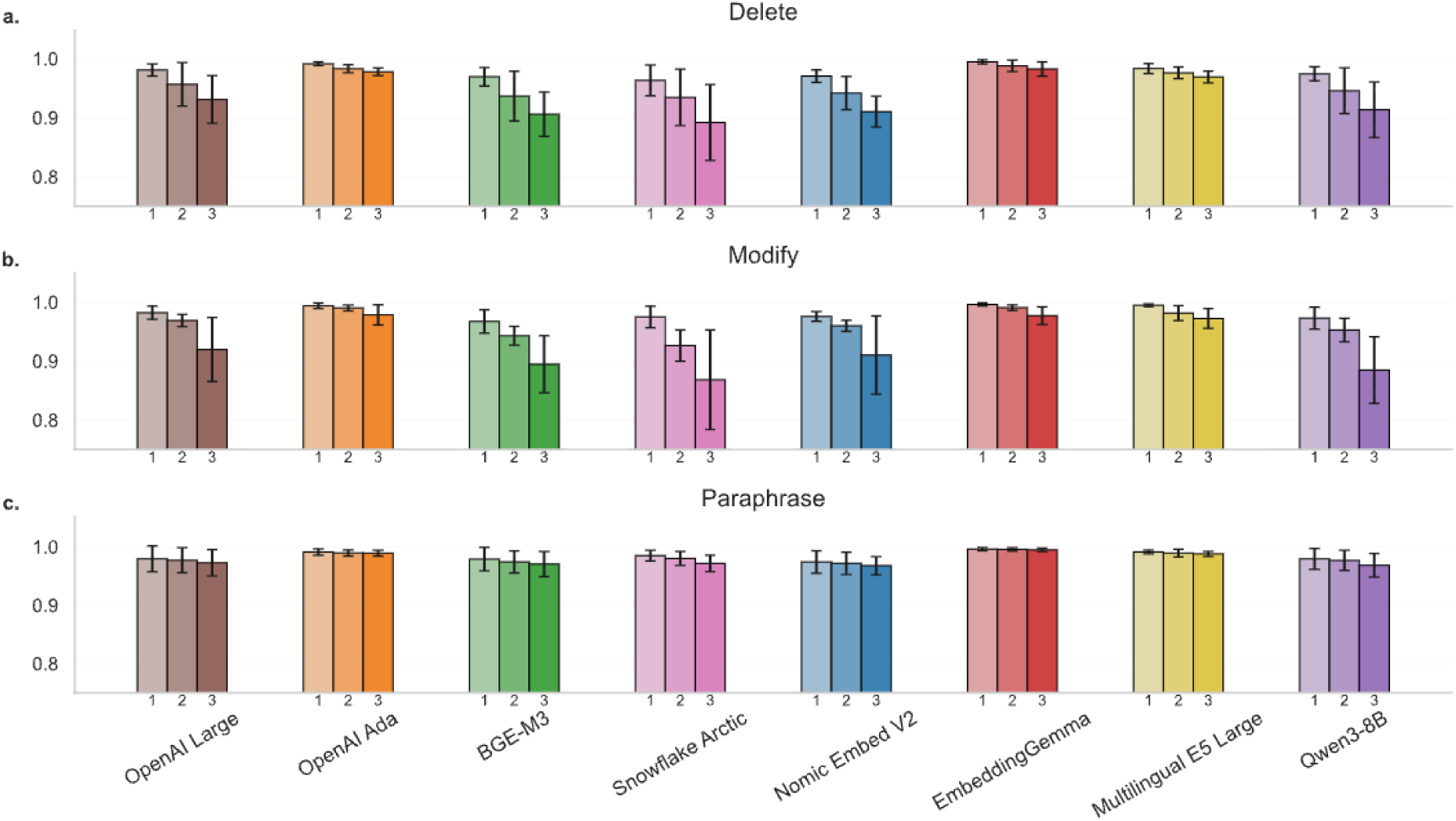
Cosine similarity between embedding vectors of Swedish summaries. The bars show average cosine similarities over the comparisons between vectors of original Swedish summaries and their perturbed versions of the same type (Delete, Modify, Paraphrase) and level (1 to 3). The black markers overlapping the bars represent standard deviations.

**Supplemental Figure S2.**
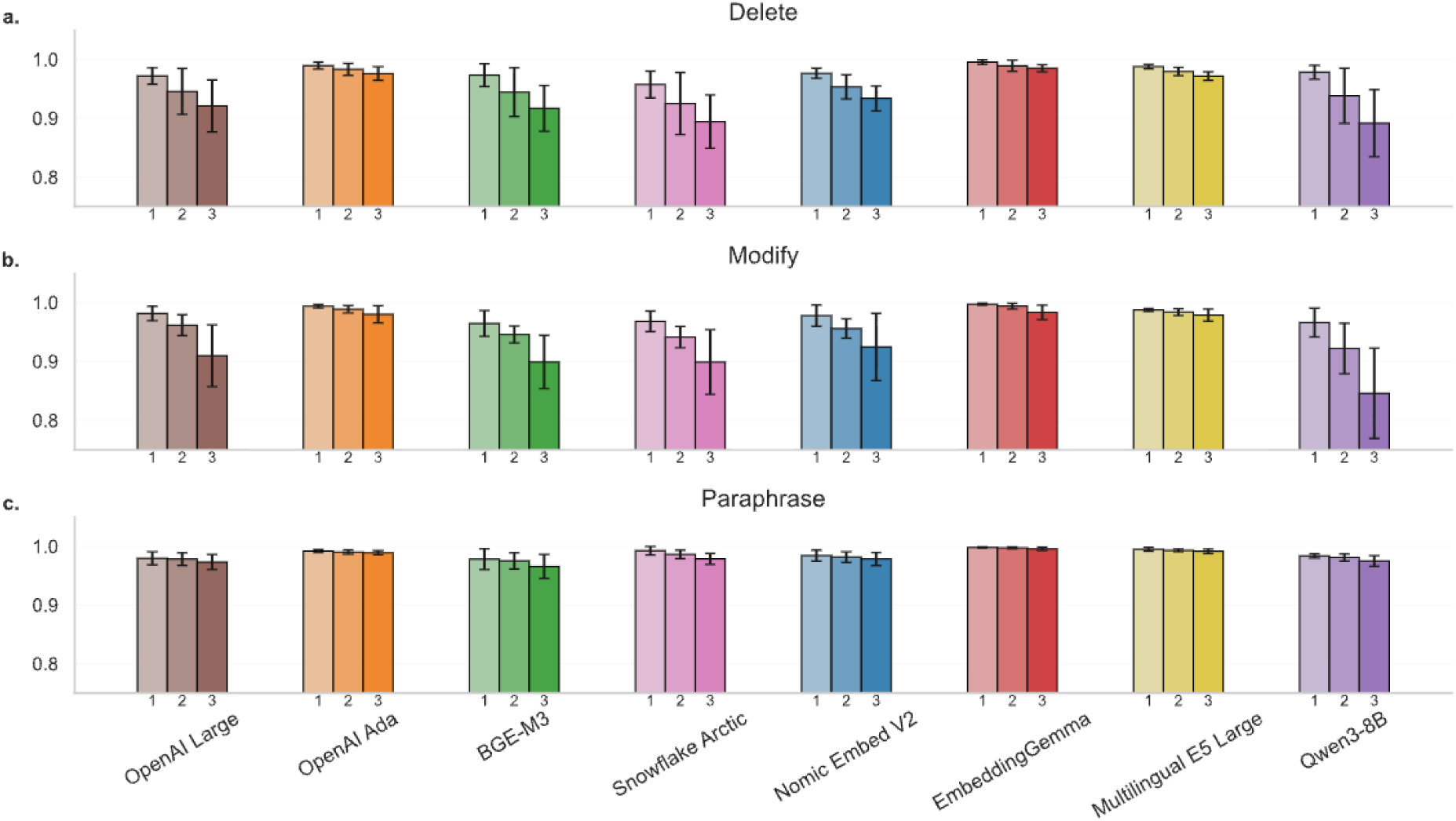
Cosine similarity between embedding vectors of Finnish summaries. The bars show average cosine similarities over the comparisons between vectors of original Finnish summaries and their perturbed versions of the same type (Delete, Modify, Paraphrase) and level (1 to 3). The black markers overlapping the bars represent standard deviations.

**Supplemental Figure S3.**
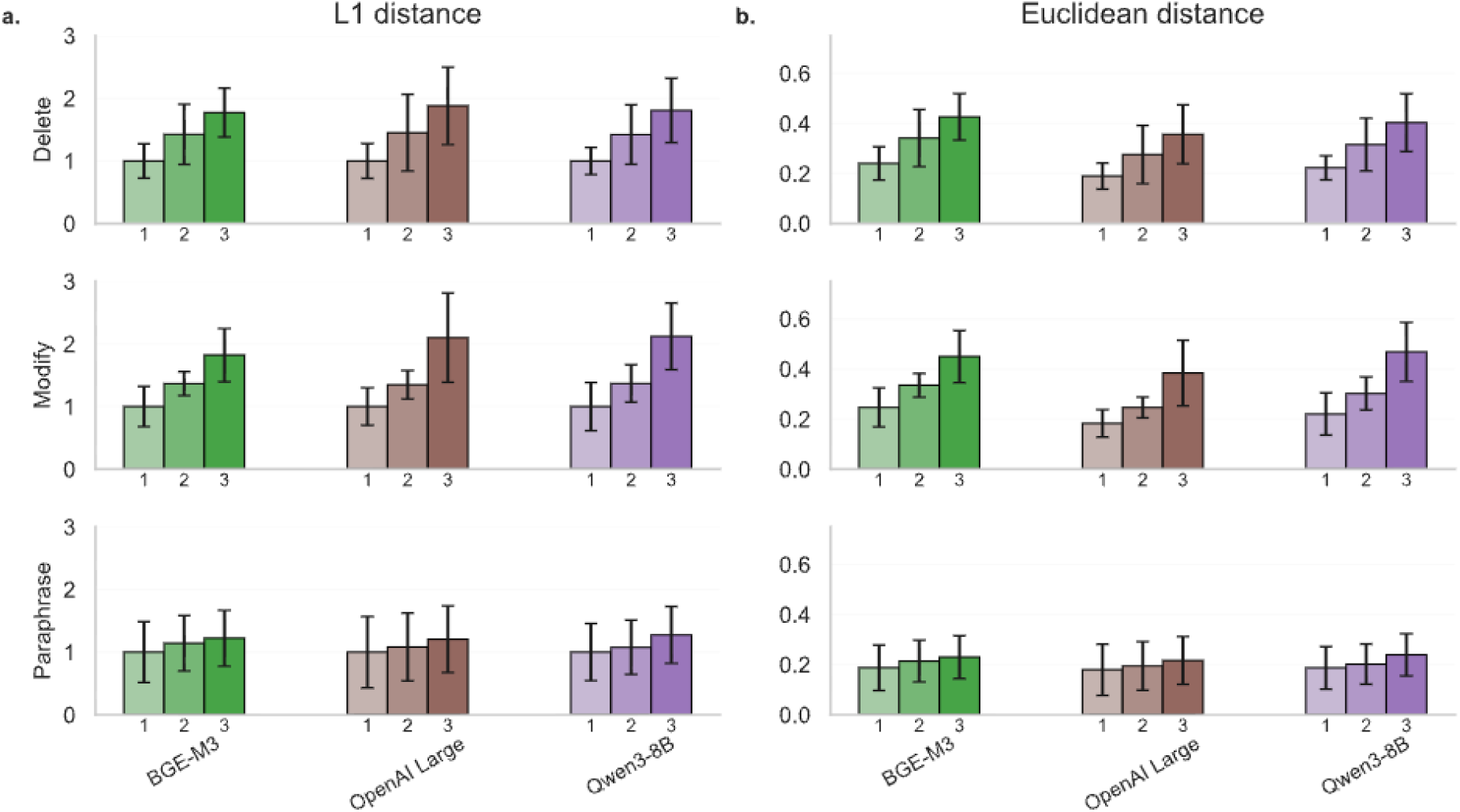
L^1^ and Euclidean distances between embedding vectors of Swedish summaries for three models. The bars show average distances between vectors of original Swedish summaries and their perturbed versions of the same type (Delete, Modify, Paraphrase) and level (1 to 3). The black markers overlapping the bars represent standard deviations. Since dimensionality affects L^1^ values strongly, they have been scaled so that level 1 value equals 1 to help comparability between models.

**Supplemental Figure S4.**
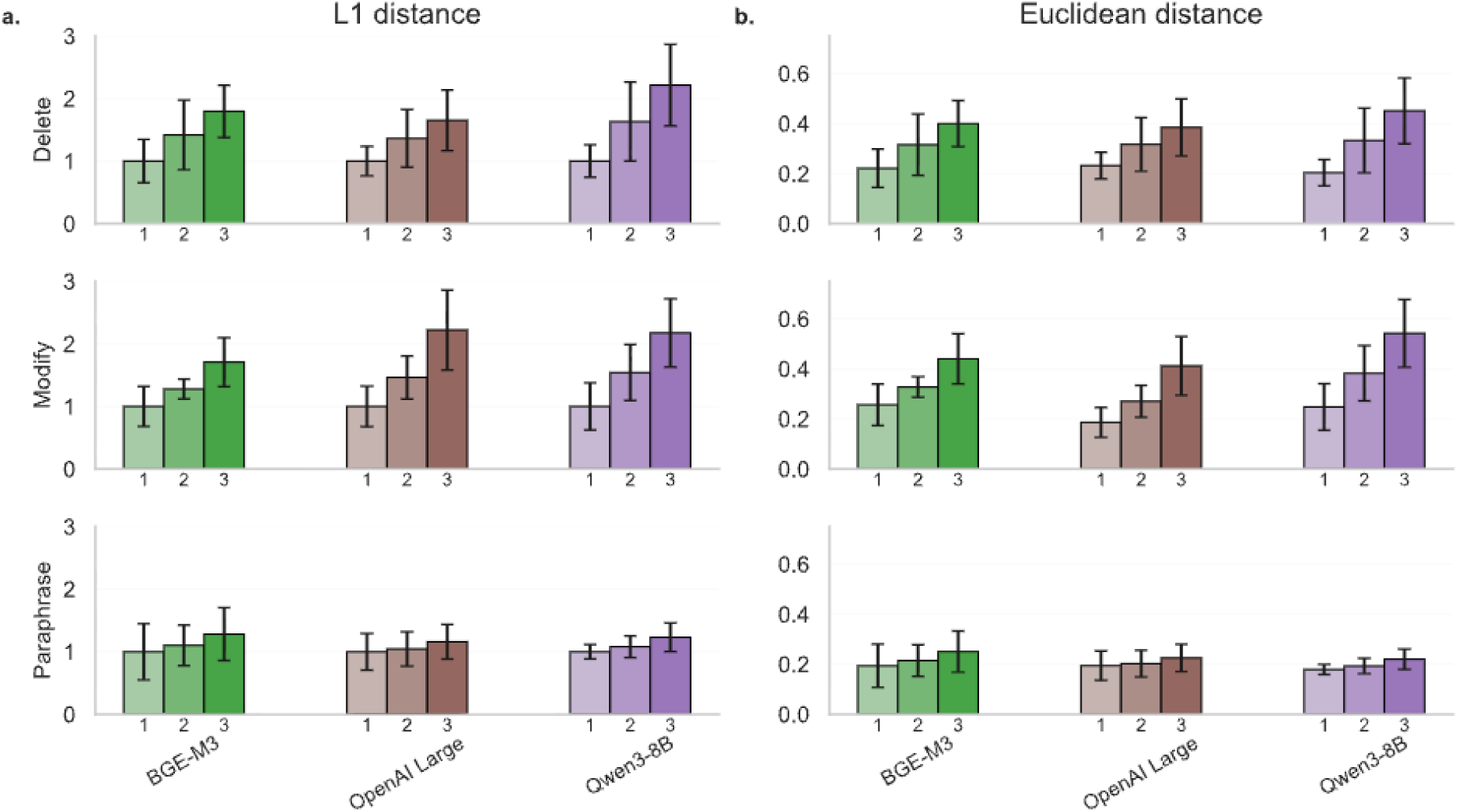
L^1^ and Euclidean distances between embedding vectors of Finnish summaries for three models. The bars show average distances between vectors of original Finnish summaries and their perturbed versions of the same type (Delete, Modify, Paraphrase) and level (1 to 3). The black markers overlapping the bars represent standard deviations. Since dimensionality affects L^1^ values strongly, they have been scaled so that level 1 value equals 1 to help comparability between models.

## Supplemental Tables

**Supplemental Table S1.** Linear regression results for the effect of perturbation level on cosine similarity, L1 distance, and Euclidean distance, grouped by language and perturbation type. Each β coefficient represents the estimated change in the respective metric per unit increase in perturbation level, with 95% confidence intervals. All models were fitted separately for each language and edit type combination.

| Language | Perturbation type | Cosine similarity $\beta$ (95% CI) | Cosine similarity p | L1 distance $\beta$ (95% CI) | L1 distance p | Euclidean distance $\beta$ (95% CI) | Euclidean distance p |
| --- | --- | --- | --- | --- | --- | --- | --- |
| English | Delete | -0.028<br>(-0.041 to -0.015) | <.001 | 2.141<br>(1.394 to 2.889) | <.001 | 0.085<br>(0.055 to 0.115) | <.001 |
| English | Modify | -0.038<br>(-0.051 to -0.025) | <.001 | 2.663<br>(1.810 to 3.515) | <.001 | 0.103<br>(0.070 to 0.137) | <.001 |
| English | Paraphrase | -0.005<br>(-0.007 to -0.002) | 0.001 | 0.566<br>(0.231 to 0.900) | <.001 | 0.022<br>(0.010 to 0.035) | <.001 |
| Swedish | Delete | -0.032<br>(-0.046 to -0.017) | <.001 | 2.342<br>(1.386 to 3.298) | <.001 | 0.093<br>(0.055 to 0.130) | <.001 |
| Swedish | Modify | -0.036<br>(-0.054 to -0.018) | <.001 | 2.558<br>(1.487 to 3.630) | <.001 | 0.102<br>(0.059 to 0.144) | <.001 |
| Swedish | Paraphrase | -0.004<br>(-0.006 to -0.002) | <.001 | 0.532<br>(0.256 to 0.808) | <.001 | 0.021<br>(0.011 to 0.031) | <.001 |
| Finnish | Delete | -0.028<br>(-0.036 to -0.020) | <.001 | 2.235<br>(1.779 to 2.691) | <.001 | 0.089<br>(0.071 to 0.107) | <.001 |
| Finnish | Modify | -0.033<br>(-0.051 to -0.014) | <.001 | 2.308<br>(1.181 to 3.436) | <.001 | 0.092<br>(0.047 to 0.137) | <.001 |
| Finnish | Paraphrase | -0.006<br>(-0.009 to -0.003) | <.001 | 0.699<br>(0.411 to 0.987) | <.001 | 0.028<br>(0.017 to 0.040) | <.001 |

**Supplemental Table S2.**
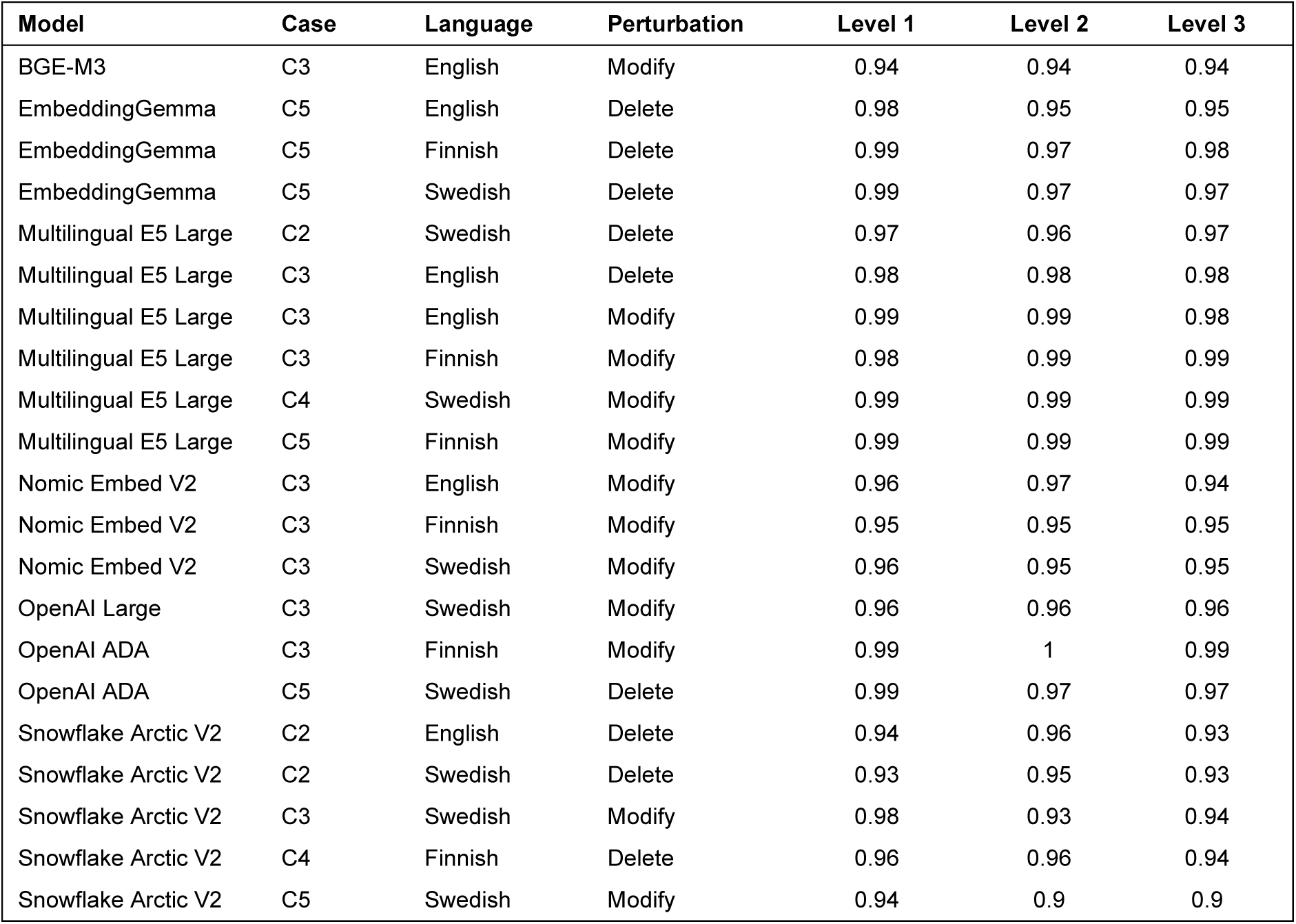
Case-level conflicting changes in cosine similarity values in perturbation types Delete and Modify. Conflicting change was defined as an increase in cosine similarity between successive perturbation levels (level 2 > level 1 or level 3 > level 2).

| Model | Case | Language | Perturbation | Level 1 | Level 2 | Level 3 |
| --- | --- | --- | --- | --- | --- | --- |
| BGE-M3 | C3 | English | Modify | 0.94 | 0.94 | 0.94 |
| EmbeddingGemma | C5 | English | Delete | 0.98 | 0.95 | 0.95 |
| EmbeddingGemma | C5 | Finnish | Delete | 0.99 | 0.97 | 0.98 |
| EmbeddingGemma | C5 | Swedish | Delete | 0.99 | 0.97 | 0.97 |
| Multilingual E5 Large | C2 | Swedish | Delete | 0.97 | 0.96 | 0.97 |
| Multilingual E5 Large | C3 | English | Delete | 0.98 | 0.98 | 0.98 |
| Multilingual E5 Large | C3 | English | Modify | 0.99 | 0.99 | 0.98 |
| Multilingual E5 Large | C3 | Finnish | Modify | 0.98 | 0.99 | 0.99 |
| Multilingual E5 Large | C4 | Swedish | Modify | 0.99 | 0.99 | 0.99 |
| Multilingual E5 Large | C5 | Finnish | Modify | 0.99 | 0.99 | 0.99 |
| Nomic Embed V2 | C3 | English | Modify | 0.96 | 0.97 | 0.94 |
| Nomic Embed V2 | C3 | Finnish | Modify | 0.95 | 0.95 | 0.95 |
| Nomic Embed V2 | C3 | Swedish | Modify | 0.96 | 0.95 | 0.95 |
| OpenAI Large | C3 | Swedish | Modify | 0.96 | 0.96 | 0.96 |
| OpenAI ADA | C3 | Finnish | Modify | 0.99 | 1 | 0.99 |
| OpenAI ADA | C5 | Swedish | Delete | 0.99 | 0.97 | 0.97 |
| Snowflake Arctic V2 | C2 | English | Delete | 0.94 | 0.96 | 0.93 |
| Snowflake Arctic V2 | C2 | Swedish | Delete | 0.93 | 0.95 | 0.93 |
| Snowflake Arctic V2 | C3 | Swedish | Modify | 0.98 | 0.93 | 0.94 |
| Snowflake Arctic V2 | C4 | Finnish | Delete | 0.96 | 0.96 | 0.94 |
| Snowflake Arctic V2 | C5 | Swedish | Modify | 0.94 | 0.9 | 0.9 |

**Supplemental Table S3.** Case-level conflicting changes in L^1^ distances in perturbation types Delete and Modify. Conflicting change was defined as a decrease in L^1^ distance between successive perturbation levels (level 1 > level 2 or level 2 > level 3).

| Model | Case | Language | Perturbation | Level 1 | Level 2 | Level 3 |
| --- | --- | --- | --- | --- | --- | --- |
| BGE-M3 | C3 | English | Modify | 8.69 | 8.79 | 8.57 |
| EmbeddingGemma | C5 | English | Delete | 3.77 | 7.01 | 6.91 |
| EmbeddingGemma | C5 | Finnish | Delete | 3.32 | 5.11 | 4.87 |
| EmbeddingGemma | C5 | Swedish | Delete | 3.11 | 5.11 | 5.05 |
| Multilingual E5 Large | C2 | Swedish | Delete | 6.32 | 7.14 | 6.61 |
| Multilingual E5 Large | C3 | English | Delete | 5.1 | 4.89 | 5.5 |
| Multilingual E5 Large | C3 | English | Modify | 3.89 | 3.14 | 5.03 |
| Multilingual E5 Large | C3 | Finnish | Modify | 4.44 | 4.32 | 4.12 |
| Multilingual E5 Large | C4 | Finnish | Modify | 4.21 | 5.55 | 5.49 |
| Multilingual E5 Large | C4 | Swedish | Modify | 2.81 | 2.73 | 4.15 |
| Multilingual E5 Large | C5 | Finnish | Modify | 4.22 | 3.28 | 3.52 |
| Nomic Embed V2 | C3 | English | Modify | 5.81 | 5.33 | 7.67 |
| Nomic Embed V2 | C3 | Finnish | Modify | 7.14 | 6.94 | 7 |
| Nomic Embed V2 | C3 | Swedish | Modify | 5.67 | 6.74 | 6.67 |
| Nomic Embed V2 | C4 | English | Modify | 3.88 | 6.67 | 6.64 |
| OpenAI Large | C3 | Swedish | Modify | 11.88 | 11.61 | 12.44 |
| OpenAI ADA | C3 | Finnish | Modify | 3.18 | 2.96 | 3.12 |
| OpenAI ADA | C4 | English | Delete | 3.27 | 3.27 | 3.99 |
| OpenAI ADA | C5 | Swedish | Delete | 4.13 | 7.36 | 7.21 |
| Snowflake Arctic V2 | C2 | English | Delete | 8.51 | 6.76 | 9.76 |
| Snowflake Arctic V2 | C2 | Swedish | Delete | 9.05 | 7.85 | 9.7 |
| Snowflake Arctic V2 | C3 | Swedish | Modify | 4.99 | 9.27 | 8.98 |
| Snowflake Arctic V2 | C4 | Finnish | Delete | 7.2 | 7.13 | 8.44 |
| Snowflake Arctic V2 | C5 | Swedish | Modify | 8.39 | 11.58 | 11.33 |

**Supplemental Table S4.**
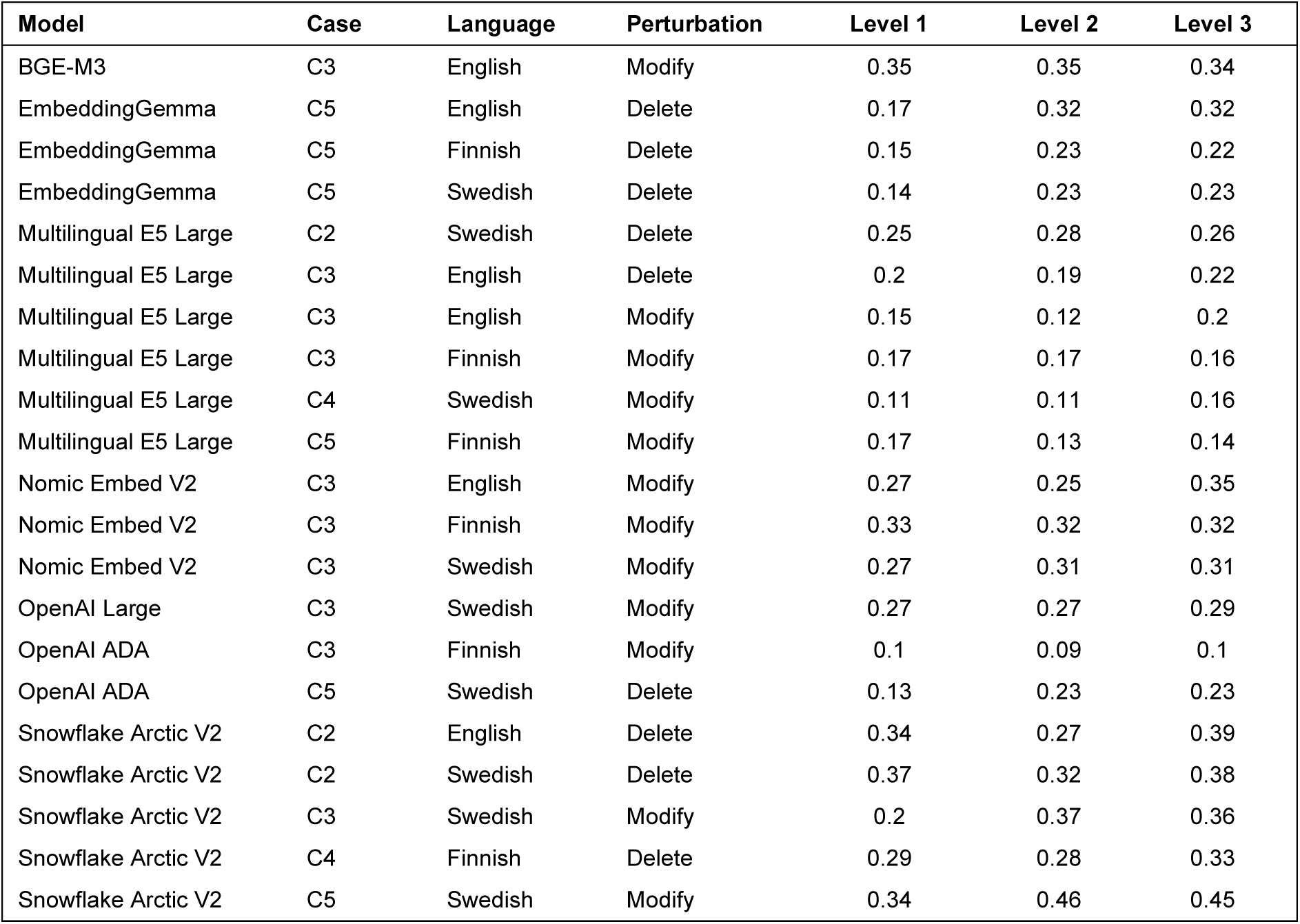
Case-level conflicting changes in Euclidean distances in perturbation types Delete and Modify. Conflicting change was defined as a decrease in Euclidean distance between successive perturbation levels (level 1 > level 2 or level 2 > level 3).

| Model | Case | Language | Perturbation | Level 1 | Level 2 | Level 3 |
| --- | --- | --- | --- | --- | --- | --- |
| BGE-M3 | C3 | English | Modify | 0.35 | 0.35 | 0.34 |
| EmbeddingGemma | C5 | English | Delete | 0.17 | 0.32 | 0.32 |
| EmbeddingGemma | C5 | Finnish | Delete | 0.15 | 0.23 | 0.22 |
| EmbeddingGemma | C5 | Swedish | Delete | 0.14 | 0.23 | 0.23 |
| Multilingual E5 Large | C2 | Swedish | Delete | 0.25 | 0.28 | 0.26 |
| Multilingual E5 Large | C3 | English | Delete | 0.2 | 0.19 | 0.22 |
| Multilingual E5 Large | C3 | English | Modify | 0.15 | 0.12 | 0.2 |
| Multilingual E5 Large | C3 | Finnish | Modify | 0.17 | 0.17 | 0.16 |
| Multilingual E5 Large | C4 | Swedish | Modify | 0.11 | 0.11 | 0.16 |
| Multilingual E5 Large | C5 | Finnish | Modify | 0.17 | 0.13 | 0.14 |
| Nomic Embed V2 | C3 | English | Modify | 0.27 | 0.25 | 0.35 |
| Nomic Embed V2 | C3 | Finnish | Modify | 0.33 | 0.32 | 0.32 |
| Nomic Embed V2 | C3 | Swedish | Modify | 0.27 | 0.31 | 0.31 |
| OpenAI Large | C3 | Swedish | Modify | 0.27 | 0.27 | 0.29 |
| OpenAI ADA | C3 | Finnish | Modify | 0.1 | 0.09 | 0.1 |
| OpenAI ADA | C5 | Swedish | Delete | 0.13 | 0.23 | 0.23 |
| Snowflake Arctic V2 | C2 | English | Delete | 0.34 | 0.27 | 0.39 |
| Snowflake Arctic V2 | C2 | Swedish | Delete | 0.37 | 0.32 | 0.38 |
| Snowflake Arctic V2 | C3 | Swedish | Modify | 0.2 | 0.37 | 0.36 |
| Snowflake Arctic V2 | C4 | Finnish | Delete | 0.29 | 0.28 | 0.33 |
| Snowflake Arctic V2 | C5 | Swedish | Modify | 0.34 | 0.46 | 0.45 |

**Supplemental Table S5.** The differences between scaled L^1^ average values with respect to full vectors and partial vectors for Qwen3-8B. The values were scaled so that Level 1 value was 1 for each triple with the same truncation type, perturbation type, and language. Using the 100 biggest absolute values instead of full vectors was enough to get us close to the original values whereas the difference was much larger using the 100 smallest absolute values.

| Model | Truncation type | Perturbation type | Language | Level 2 difference (%) | Level 3 difference (%) |
| --- | --- | --- | --- | --- | --- |
| Qwen3-8B | max100 | Delete | English | -1.1 | 1.5 |
| Qwen3-8B | max100 | Delete | Finnish | 0.4 | 0.1 |
| Qwen3-8B | max100 | Delete | Swedish | -0.1 | 0.6 |
| Qwen3-8B | max100 | Modify | English | 1.7 | 1.7 |
| Qwen3-8B | max100 | Modify | Finnish | -0.0 | 1.7 |
| Qwen3-8B | max100 | Modify | Swedish | 1.7 | 0.1 |
| Qwen3-8B | max100 | Paraphrase | English | -0.2 | 0.3 |
| Qwen3-8B | max100 | Paraphrase | Finnish | -1.3 | -1.7 |
| Qwen3-8B | max100 | Paraphrase | Swedish | -0.0 | -0.1 |
| Qwen3-8B | first100 | Delete | English | 3.3 | 4.3 |
| Qwen3-8B | first100 | Delete | Finnish | 4.0 | 5.3 |
| Qwen3-8B | first100 | Delete | Swedish | -2.4 | -0.1 |
| Qwen3-8B | first100 | Modify | English | -3.6 | 2.0 |
| Qwen3-8B | first100 | Modify | Finnish | 3.0 | 4.7 |
| Qwen3-8B | first100 | Modify | Swedish | 0.4 | 2.1 |
| Qwen3-8B | first100 | Paraphrase | English | 2.4 | 0.2 |
| Qwen3-8B | first100 | Paraphrase | Finnish | 0.3 | -0.8 |
| Qwen3-8B | first100 | Paraphrase | Swedish | -2.1 | -0.1 |
| Qwen3-8B | min100 | Delete | English | -1.5 | -0.3 |
| Qwen3-8B | min100 | Delete | Finnish | 1.7 | 6.3 |
| Qwen3-8B | min100 | Delete | Swedish | 7.9 | -0.4 |
| Qwen3-8B | min100 | Modify | English | -1.0 | 7.9 |
| Qwen3-8B | min100 | Modify | Finnish | 4.6 | -10.4 |
| Qwen3-8B | min100 | Modify | Swedish | 0.5 | -10.8 |
| Qwen3-8B | min100 | Paraphrase | English | -5.5 | 7.0 |
| Qwen3-8B | min100 | Paraphrase | Finnish | 3.4 | -4.8 |
| Qwen3-8B | min100 | Paraphrase | Swedish | 3.4 | 6.4 |

**Supplemental Table S6.** The differences between scaled L^1^ average values with respect to full vectors and partial vectors for EmbeddingGemma. The values were scaled so that Level 1 value was 1 for each triple with the same truncation type, perturbation type, and language. Using the 100 biggest absolute values instead of full vectors was enough to get us close to the original values whereas the difference was much larger using the 100 smallest absolute values.

| Model | Truncation type | Perturbation type | Language | Level 2 difference (%) | Level 3 difference (%) |
| --- | --- | --- | --- | --- | --- |
| EmbeddingGemma | max100 | Delete | English | -0.7 | 0.1 |
| EmbeddingGemma | max100 | Delete | Finnish | 0.0 | -0.5 |
| EmbeddingGemma | max100 | Delete | Swedish | 0.2 | 1.2 |
| EmbeddingGemma | max100 | Modify | English | 3.2 | 2.3 |
| EmbeddingGemma | max100 | Modify | Finnish | -0.8 | -0.6 |
| EmbeddingGemma | max100 | Modify | Swedish | -1.0 | -1.2 |
| EmbeddingGemma | max100 | Paraphrase | English | 0.7 | -1.5 |
| EmbeddingGemma | max100 | Paraphrase | Finnish | -1.0 | -0.4 |
| EmbeddingGemma | max100 | Paraphrase | Swedish | -1.0 | -1.8 |
| EmbeddingGemma | first100 | Delete | English | -1.7 | 0.2 |
| EmbeddingGemma | first100 | Delete | Finnish | 2.0 | 5.4 |
| EmbeddingGemma | first100 | Delete | Swedish | -0.8 | -0.3 |
| EmbeddingGemma | first100 | Modify | English | -0.5 | 5.7 |
| EmbeddingGemma | first100 | Modify | Finnish | -0.6 | -1.3 |
| EmbeddingGemma | first100 | Modify | Swedish | 0.5 | 0.2 |
| EmbeddingGemma | first100 | Paraphrase | English | 3.2 | -1.3 |
| EmbeddingGemma | first100 | Paraphrase | Finnish | -0.3 | 7.3 |
| EmbeddingGemma | first100 | Paraphrase | Swedish | -1.3 | 1.3 |
| EmbeddingGemma | min100 | Delete | English | -0.8 | 4.1 |
| EmbeddingGemma | min100 | Delete | Finnish | -8.4 | -2.7 |
| EmbeddingGemma | min100 | Delete | Swedish | 6.9 | -4.2 |
| EmbeddingGemma | min100 | Modify | English | 2.8 | -2.6 |
| EmbeddingGemma | min100 | Modify | Finnish | 3.0 | -0.4 |
| EmbeddingGemma | min100 | Modify | Swedish | 2.4 | -6.0 |
| EmbeddingGemma | min100 | Paraphrase | English | 4.4 | 12.5 |
| EmbeddingGemma | min100 | Paraphrase | Finnish | 2.9 | 5.2 |
| EmbeddingGemma | min100 | Paraphrase | Swedish | 17.8 | 4.1 |

**Supplemental Table S7.** Dimensions that never appeared among the 100 biggest changes. We computed the difference vectors and recorded where the 100 biggest changes happened. The number of dimensions that never appeared among the 100 biggest changes increased for both models from multilingual to monolingual texts and from all perturbation types to a single perturbation type.

| Model | Perturbation type | Language | Number of vectors | Dimensions outside top 100 (%) |
| --- | --- | --- | --- | --- |
| Qwen3-8B | All | All | 135 | 19.9 |
| Qwen3-8B | All | English | 45 | 47.7 |
| Qwen3-8B | All | Finnish | 45 | 49.5 |
| Qwen3-8B | All | Swedish | 45 | 50.2 |
| Qwen3-8B | Delete | All | 45 | 51.3 |
| Qwen3-8B | Delete | English | 15 | 76.1 |
| Qwen3-8B | Delete | Finnish | 15 | 74.7 |
| Qwen3-8B | Delete | Swedish | 15 | 76.2 |
| Qwen3-8B | Modify | All | 45 | 53.2 |
| Qwen3-8B | Modify | English | 15 | 75.9 |
| Qwen3-8B | Modify | Finnish | 15 | 75.6 |
| Qwen3-8B | Modify | Swedish | 15 | 76.7 |
| Qwen3-8B | Paraphrase | All | 45 | 52.2 |
| Qwen3-8B | Paraphrase | English | 15 | 77.7 |
| Qwen3-8B | Paraphrase | Finnish | 15 | 81.1 |
| Qwen3-8B | Paraphrase | Swedish | 15 | 78.8 |
| EmbeddingGemma | All | All | 135 | 0.4 |
| EmbeddingGemma | All | English | 45 | 4.7 |
| EmbeddingGemma | All | Finnish | 45 | 4.0 |
| EmbeddingGemma | All | Swedish | 45 | 4.3 |
| EmbeddingGemma | Delete | All | 45 | 4.8 |
| EmbeddingGemma | Delete | English | 15 | 26.7 |
| EmbeddingGemma | Delete | Finnish | 15 | 25.7 |
| EmbeddingGemma | Delete | Swedish | 15 | 26.2 |
| EmbeddingGemma | Modify | All | 45 | 4.6 |
| EmbeddingGemma | Modify | English | 15 | 25.4 |
| EmbeddingGemma | Modify | Finnish | 15 | 24.2 |
| EmbeddingGemma | Modify | Swedish | 15 | 28.8 |
| EmbeddingGemma | Paraphrase | All | 45 | 4.6 |
| EmbeddingGemma | Paraphrase | English | 15 | 32.8 |
| EmbeddingGemma | Paraphrase | Finnish | 15 | 31.1 |
| EmbeddingGemma | Paraphrase | Swedish | 15 | 29.8 |

